# Characteristics and Predictors of Apparent Treatment Resistant Hypertension in Real-World Populations Using Electronic Health Record-Based Data

**DOI:** 10.1101/2023.04.28.23289293

**Authors:** Eissa Jafari, Rhonda M. Cooper-DeHoff, Mark B. Effron, William R. Hogan, Caitrin W. McDonough

## Abstract

**Background:** Apparent treatment-resistant hypertension (aTRH) is defined as uncontrolled blood pressure (BP) despite using ≥3 antihypertensive classes or controlled BP while using ≥4 antihypertensive classes. Patients with aTRH have a higher risk for adverse cardiovascular outcomes compared to patients with controlled hypertension. Although there have been prior reports on the prevalence, characteristics, and predictors of aTRH, these have been broadly derived from smaller datasets, randomized controlled trials, or closed healthcare systems.

**Methods:** We extracted patients with hypertension defined by ICD 9 and 10 codes during 1/1/2015-12/31/2018, from two large electronic health record databases: the OneFlorida Data Trust (n=223,384) and Research Action for Health Network (REACHnet) (n=175,229). We applied our previously validated aTRH and stable controlled hypertension (HTN) computable phenotype algorithms and performed univariate and multivariate analyses to identify the prevalence, characteristics, and predictors of aTRH in these real-world populations.

**Results:** The prevalence of aTRH in OneFlorida (16.7%) and REACHnet (11.3%) was similar to prior reports. Both populations had a significantly higher proportion of black patients with aTRH compared to those with stable controlled HTN. aTRH in both populations shared similar significant predictors, including black race, diabetes, heart failure, chronic kidney disease, cardiomegaly, and higher body mass index. In both populations, aTRH was significantly associated with similar comorbidities, when compared with stable controlled HTN.

**Conclusion:** In two large, diverse real-world populations, we observed similar comorbidities and predictors of aTRH as prior studies. In the future, these results may be used to improve healthcare professionals’ understanding of aTRH predictors and associated comorbidities.

**Clinical Perspective:** *What Is New?:*

- Prior studies of apparent treatment resistant hypertension have focused on cohorts from smaller datasets, randomized controlled trials, or closed healthcare systems.
- We used validated computable phenotype algorithms for apparent treatment resistant hypertension and stable controlled hypertension to identify the prevalence, characteristics, and predictors of apparent treatment resistant hypertension in two large, diverse real-world populations.

*What Are the Clinical Implications?:*

- Large, diverse real-world populations showed a similar prevalence of aTRH, 16.7% in OneFlorida and 11.3% in REACHnet, compared to those observed from other cohorts.
- Patients classified as apparent treatment resistant hypertension were significantly older and had a higher prevalence of comorbid conditions such as diabetes, dyslipidemia, coronary artery disease, heart failure with preserved ejection fraction, and chronic kidney disease stages 1-3.
- Within diverse, real-world populations, the strongest predictors for apparent treatment resistant hypertension were black race, higher body mass index, heart failure, chronic kidney disease, and diabetes.

## Introduction

Hypertension (HTN) is a major modifiable risk factor for cardiovascular morbidity and mortality [1]. Based on the blood pressure (BP) cut-off (≤ 130/80 mm Hg), recommended by the 2017 American College of Cardiology/American Heart Association (ACC/AHA) clinical practice guideline, HTN affects ∼121.5 million adults in the United States (U.S), almost half of the US adult population [2]. While tailored antihypertensive medications substantially control HTN, a considerable proportion of patients with HTN fail to achieve their target BP [3]. Resistant hypertension (RHTN) is defined as uncontrolled BP despite using ≥3 antihypertensive medications from different classes or controlled BP while using ≥4 antihypertensive medications from different classes. Patients classified as having RHTN based solely on medication prescription information and/or BP information can be controlled, uncontrolled, pseudo-resistant, or refractory hypertension patients. Thus, apparent treatment-resistant hypertension (aTRH) is the commonly used term for epidemiological studies when pseudo-resistant hypertension and medication adherence cannot be determined [4].

Analyses of population and clinic-based data suggest that aTRH is prevalent in 4.3 - 29.7% of patients with treated HTN [5]. Though aTRH prevalence is widely variable, a series of studies have identified common aTRH predictors (e.g. heart failure, diabetes, black race, higher body mass index (BMI), male sex, chronic kidney disease (CKD), and older age) [6-9]. Importantly, patients with aTRH are at a higher risk for cardiovascular events and target organ damage, compared to those with controlled BP [6-8, 10-12]. This increased risk in patients with aTRH is more likely due to uncontrolled BP and concomitant comorbidities, like obesity, diabetes, CKD, left ventricular hypertrophy, obstructive sleep apnea, and others [8, 13-15]. Furthermore, aTRH is associated with worse outcomes in patients with pre-existing comorbidities. Studies have shown that aTRH is associated with a higher rate of adverse cardiovascular outcomes (e.g., stroke, myocardial infarction (MI), and death) in patients with CKD or ischemic heart diseases [6, 16]. Notably, although BP control is associated with a reduction in cardiovascular disease (CVD), lowering BP appears less protective against CVD in patients with aTRH [17]. Collectively, these findings necessitate the need for early identification of those at risk for aTRH, as they would benefit from specific therapeutic interventions.

Our current understanding of aTRH characteristics and predictors is derived from randomized controlled trials (RCTs), small real-world cohorts, or closed health systems, predominantly of European ancestry. Hence, leveraging our validated aTRH computable phenotype algorithm [18], we aimed to identify characteristics and predictors of aTRH within electronic health records (EHRs)-based data of large, racially and ethnically diverse populations from the OneFlorida Data Trust and the Research Action for Health Network (REACHnet). We used OneFlorida as a discovery population and REACHnet as a validation cohort. We hypothesized that the large and diverse patient populations in OneFlorida and REACHnet would enable identifying and validating real-world characteristics and predictors of aTRH.

## Methods

### Data Source

A retrospective observational cohort study was conducted on EHR-based data from ambulatory HTN patients, collected from the OneFlorida Data Trust and REACHnet, from January 1^st^, 2015, through December 31^st^, 2018. Coordinated by the OneFlorida Clinical Research Consortium, the OneFlorida Data Trust is a patient-centered research data repository, comprising EHR-based data, claims-based data, and other data on > 17 million Floridians, from ten healthcare systems across the state of Florida [19]. REACHnet is managed by the Louisiana Public Health Institute (LPHI) and houses clinical records on > 8 million individuals from different healthcare systems and academic centers in Louisiana and Eastern Texas [20]. OneFlorida and REACHnet have been part of the National Patient-Centered Clinical Research Network (PCORnet), and data from OneFlorida and REACHnet are stored in the PCORnet common data model (CDM). [21]. From OneFlorida, data were included from all sites. From REACHnet, data from Ochsner Health were included.

### Patient Population

Patients were included in the study if they had the following criteria: adults ≥ 18 years old, a diagnosis of HTN in an ambulatory setting based on the *International Classification of Diseases, Ninth Revision* (ICD-9: 401.0, 401.1, 401.9) or *Tenth Revision* (ICD-10: I10) codes, at least two BP measurements where systolic and diastolic BP are not null, and at least one prescribing record. Patients with secondary HTN, pregnancy, and missing drug exposure and BP data were excluded from the analysis. The study was approved by the institutional review boards at the University of Florida and the Ochsner Clinic Foundation, with a full waiver of informed consent for research involving data previously collected for non-research purposes.

### Data Fields and Extraction

Data were extracted in the PCORnet CDM format on 8/16/2019 from the OneFlorida Data Trust, and on 5/3/2022 from REACHnet. The following tables from the PCORnet CDM were included: demographic, condition, diagnosis, encounter, procedures, prescribing, and vital.

### aTRH and Stable Controlled HTN Computable Phenotype Algorithms

We used our previously validated computable phenotype algorithms to classify patients as aTRH, stable controlled HTN, or other HTN in the OneFlorida and REACHnet datasets [18]. Briefly, adult patients (≥18 years) with an outpatient HTN diagnosis, prescription records, and BP measurements during the study period were included in the data preparation steps. First, we used our optimized antihypertensive medications classification system to map all prescribed medications in the HTN populations to antihypertensive classes using RxNorm Concept Unique Identifiers (RxCUI) [18, 22]. Second, we created a medication exposure variable to capture the number of daily antihypertensive drug classes every day, over the study period. Lastly, systolic, and diastolic BP measurements were considered during the antihypertensive medication prescribing windows to determine BP control based on a <140/90 mm Hg cut-off [18]. This cutoff was selected because most of our data were historical, collected from January 1^st^, 2015, through December 31^st^, 2018.

Patients were classified as aTRH if they had either four concurrent antihypertensive prescriptions from different antihypertensive drug classes, regardless of BP, or three concurrent antihypertensive prescriptions from different antihypertensive drug classes while BP was ≥ 140/90 mm Hg, at least one month following the initiation of the third antihypertensive prescription. Patients were required to meet these criteria twice, at least 30 days apart but within 3 years of each other, over the study period.

Patients were classified as stable controlled HTN if they met the following criteria over the study period: one or two concurrent antihypertensive prescriptions from different antihypertensive drug classes and at least 80% of BP readings from routine outpatient care encounters classified as controlled (BP<140/90 mm Hg) during the active prescription period. Current Procedural Codes (CPT) codes were used to indicate routine outpatient care.

Patients who did not meet the criteria for aTRH or controlled HTN were classified as other HTN and included those with uncontrolled BP while prescribed one or two concurrent antihypertensive medications, or controlled BP while prescribed three concurrent antihypertensive medications [18].

For all included patients, the index date was defined as the first date the patient met the classification for aTRH or stable controlled HTN, or the second outpatient BP measurement date in the study period. Additionally, comorbidities were defined by ICD-9 and ICD-10 codes as present if the code was in the patients’ EHR at or before the index date (Supplemental Table 1). Drug exposure and average BP metrics were determined at the index date. To minimize white coat HTN, the lowest BP value was used in case the patient had multiple BP measurements at a single encounter. Antihypertensive drug classes included in the analysis were calcium channel blockers (CCBs), angiotensin-converting enzyme inhibitors (ACEIs), Angiotensin receptor blockers (ARBs), β-Blockers (BBs), thiazide-like diuretics, aldosterone antagonist, other diuretics (e.g. potassium-sparing, loop), and other antihypertensives (e.g. vasodilators, renin inhibitors, centrally acting alpha agonists, and alpha-blockers) (Supplemental Table 2).

### Statistical Analysis

Patients’ characteristics, drug exposure, and BP control were analyzed using descriptive statistics. Continuous variables were presented as mean and standard deviation (SD), whereas categorical variables were presented as absolute frequencies and percentages. Differences in patients’ characteristics and comorbidities between aTRH and stable controlled HTN cohorts were tested using the Chi-square test for categorical variables and analysis of variance (ANOVA) for quantitative variables. In addition, univariate logistic regression was performed to investigate the relationship between aTRH and independent predictors using the estimated odds ratio (OR) and 95% confidence intervals (CIs). To further determine the most significant predictors for developing aTRH, a stepwise logistic regression model was used with a cut-off of *P* < 0.05 for a variable to enter the model and a threshold of *P* < 0.001 to stay in the model. All statistical analyses were conducted by data source (OneFlorida or REACHnet), and performed using SAS version 9.4 (SAS Institute Inc., Cary, North Carolina). Results with *P* -value <0.05 were considered statistically significant. Significant predictors for aTRH with an OR ≥1.3 were selected as top predictors.

## Results

### Prevalence of aTRH and stable controlled HTN, and Patient Characteristics in OneFlorida

Of 736,158 patients identified with an outpatient HTN diagnosis within OneFlorida in the study period, 506,246 did not have BP or drug data; thus, they were excluded from the study. We also excluded 6,528 individuals with a diagnosis of secondary HTN or evidence of pregnancy, leaving 223,384 patients in the study analysis. Of those, 37,274 (16.7%) patients were identified with aTRH, whereas 32,806 (14.7%) met the criteria for stable controlled HTN (Supplemental Figure 1).

Baseline characteristics presented by HTN group are shown in Table 1. Patients with aTRH were older, more likely to be black, and had a higher proportion of BMI ≥25.0 kg/m^2^, compared with patients in the stable controlled HTN group (Table 1). Additionally, at the index date, patients with aTRH had a higher prevalence of comorbidities, including diabetes, dyslipidemia, coronary artery disease, stroke, heart failure with preserved ejection fraction (HFpEF), heart failure with reduced ejection fraction (HFrEF), myocardial infarction, CKD (stage 1-3, and stage 4-5), sleep apnea, and/or others, compared to those with stable controlled HTN (Table 1).

**Table 1.** Baseline Demographics and Characteristics of Adults with HTN, aTRH, and Stable Controlled HTN in the OneFlorida, and REACHnet populations, January 1, 2015 – December 31, 2018

| Population | OneFlorida |  |  |  | REACHnet |  |  |  |
| --- | --- | --- | --- | --- | --- | --- | --- | --- |
| Characteristic | Overall HTN | aTRH | Stable Controlled HTN | P-value | Overall HTN | aTRH | Stable Controlled HTN | P-value |
| n (%) | 223,384 (100%) | 37,274 (16.7%) | 32,806 (14.7%) |  | 175,229 (100%) | 19,863 (11,3%) | 42,128 (24%) |  |
| Age, years, mean (St. d.) | 63 ± 14 | 66 ± 13 | 62 ± 14 | <.0001 | 61.3 ± 14.1 | 61.4 ± 13.3 | 63.6 ± 12.0 | <.0001 |
| Sex (male) | 101,853 (46%) | 17,052 (46%) | 14,887 (45%) | 0.2554 | 79,875 (45%) | 8,748 (44%) | 18,770 (44%) | 0.2303 |
| Race |  |  |  |  |  |  |  |  |
| White | 146 264 (65%) | 20,594 (55%) | 23,736 (72%) |  | 112,346 (64%) | 10,196 (51%) | 29,575 (70%) |  |
| Black | 58,928 (26%) | 14,441 (39%) | 6,377 (19%) | <.0001 | 59,350 (34%) | 9,429 (47%) | 11,684 (27%) | <.0001 |
| Other* | 12,391 (6%) | 1,533 (4%) | 1,958 (6%) |  | 3,074 (1%) | 186 (<1%) | 738 (1%) |  |
| Unknown/No Information | 5,801 (3%) | 706 (2%) | 735 (2%) |  | 459 (<1%) | 52 (<1%) | 131 (<1%) |  |
| Ethnicity |  |  |  |  |  |  |  |  |
| Hispanic | 33,417 (15%) | 5,393 (15%) | 5,613 (17%) | <.0001 | 4,407 (2%) | 367 (1%) | 1,122 (2%) | <.0001 |
| Non-Hispanic | 185,188 (83%) | 31,351 (84%) | 26,685 (81%) |  | 169,972, (97%) | 19,419 (97%) | 40,871 (97%) |  |
| Unknown/No Information | 4,779 (2%) | 338 (1%) | 696 (2%) |  | 850 (<1%) | 135 (<1%) | 77 (<1%) |  |
| Body Mass Index, kg/m<sup>2</sup> |  |  |  |  |  |  |  |  |
| <25.0 | 47,983 (22%) | 6,461 (18%) | 7,808 (24%) |  | 33,417 (19%) | 2,797 (13%) | 7,903 (18%) |  |
| 25.0 to <30.0 | 67,018 (30%) | 10,132 (28%) | 10,445 (32%) | <.0001 | 51,536 (29%) | 5,000 (25%) | 13,013 (31%) | <.0001 |
| ≥30.0 | 105,731 (48%) | 20,230 (55%) | 14,393 (44%) |  | 89,456 (51%) | 12,013 (60%) | 21047 (50%) |  |
| Smoking/Tobacco Use |  |  |  |  |  |  |  |  |
| Smoker/Tobacco User | 32,486 (15%) | 4,801 (13%) | 4,220 (13%) | 0.9472 | 31,482 (18%) | 3,007 (15%) | 6,594 (15%) | 0.0991 |
| Former Smoker/Tobacco User | 71,709 (33%) | 13,908 (37%) | 11,226 (34%) | <.0001 | 33,160 (19%) | 4,458 (22%) | 8,814 (20%) | <.0001 |
| Non-Smoker/Tobacco User | 112,599 (52%) | 18,272 (49%) | 17,144 (52%) |  | 110,582 (63%) | 12,398 (62%) | 26027 (63%) |  |
| Comorbidities |  |  |  |  |  |  |  |  |
| Diabetes | 73,887 (33%) | 18,545 (50%) | 11,306 (35%) | <.0001 | 46,136 (26%) | 9,327 (46%) | 13,725 (32%) | <.0001 |
| Dyslipidemia | 121,353 (54%) | 25,879 (70%) | 21,154 (64%) | <.0001 | 77,182 (44%) | 14,070 (70%) | 27,825 (66%) | <.0001 |
| Coronary Artery Disease | 43,005 (19%) | 11,527 (31%) | 6,550 (20%) | <.0001 | 23,914 (13%) | 5,404 (27%) | 8,186 (19%) | <.0001 |
| Peripheral arterial disease | 18,109 (8%) | 5,302 (14%) | 2,600 (8%) | <.0001 | 8,247 (4%) | 2318 (11%) | 2,825 (6%) | <.0001 |
| Angina | 20,072 (9%) | 5,634 (15%) | 3,321 (10%) | <.0001 | 3170 (1%) | 1,003 (5%) | 1,295 (3%) | <.0001 |
| Myocardial Infarction | 16,203 (7%) | 4,583 (12%) | 2,537 (8%) | <.0001 | 7,500 (4%) | 1,848 (9%) | 2,614 (6%) | <.0001 |
| Stroke | 11,318 (5%) | 3,264 (9%) | 1,614 (5%) | <.0001 | 5,107 (3%) | 1,137 (5%) | 1,535 (5%) | <.0001 |
| HFrEF (Systolic HF) | 8,530 (4%) | 3,922 (11%) | 1,192 (4%) | <.0001 | 5,095 (3%) | 1,678 (8%) | 1,831 (4%) | <.0001 |
| HFpEF (All other HF) | 22,920 (10%) | 8,573 (23%) | 3,094 (9%) | <.0001 | 13,064 (7%) | 4,245 (21%) | 4,098 (9%) | <.0001 |
| Pulmonary Hypertension | 9,440 (4%) | 3,641 (10%) | 1,409 (4%) | <.0001 | 4,014 (2%) | 1,399 (7%) | 1,368 (3%) | <.0001 |
| Chronic kidney disease (stages 1-3) | 25,901 (12%) | 9,289 (25%) | 3,659 (11%) | <.0001 | 16,934 (9%) | 5,103 (25%) | 5,658 (13%) | <.0001 |
| Chronic kidney disease (stages 4-5) | 7,639 (3%) | 2,871 (8%) | 847 (3%) | <.0001 | 3,755 (2%) | 1,087 (5%) | 791 (1%) | <.0001 |
| Sleep Apnea | 27,798 (12%) | 7,416 (20%) | 4,973 (15%) | <.0001 | 12,661 (7%) | 3,468 (17%) | 5,295 (12%) | <.0001 |

Others: American Indian or Alaska Native, Asian, Native Hawaiian or Other Pacific Islander, Multiple Race.
HTN: hypertension, aTRH: apparent treatment resistant hypertension, HFrEF: heart failure with reduced ejection fraction, HFpEF: heart failure with preserved ejection fraction.

### Prescribed Antihypertensive Medications and BP Metrics in OneFlorida

On average, aTRH and stable controlled HTN patients were taking 3.7 ± 0.6 and 1.4 ± 0.5 antihypertensive classes, respectively (Table 2). The most frequently prescribed antihypertensive classes at the index date among the aTRH cohort were BBs (77%), CCBs (71%), and thiazide-like diuretics (57%). In those with stable controlled HTN, ACEIs (39%), BBs (31%), and thiazide-like diuretics (23%) were the most commonly prescribed classes (Table 2).

**Table 2.** Antihypertensive Medications Prescribed at Index Date for Adults with HTN, aTRH, and stable controlled HTN in the OneFlorida, and REACHnet populations.

| **Population** | **OneFlorida** |  |  | **REACHnet** |  |  |
| --- | --- | --- | --- | --- | --- | --- |
| **Antihypertensive Medications** | **Overall**<br>**N=223,384** | **aTRH**<br>**N=37,274** | **Stable**<br>**Controlled**<br>**HTN**<br>**N=32,806** | **Overall HTN**<br>**175,229** | **aTRH**<br>**19,863** | **Stable**<br>**Controlled**<br>**HTN**<br>**42,128** |
| **Medication Class** |  |  |  |  |  |  |
| Calcium channel blockers | 62,127 (28%) | 26,563 (71%) | 6,040 (18%) | 35,797 (20%) | 14,754 (74%) | 8,508 (20%) |
| Angiotensin-converting enzyme (ACE) inhibitors | 75,398 (34%) | 19,966 (54%) | 12,757 (39%) | 40,775 (23%) | 9,323 (47%) | 14,948 (35%) |
| Angiotensin receptor blockers (ARBs) | 43,635 (20%) | 16,114 (43%) | 5,521 (17%) | 26,492 (15%) | 9,039 (46%) | 8,288 (20%) |
| β-Blockers | 72,289 (32%) | 28,557 (77%) | 10,292 (31%) | 36,150 (21%) | 14,895 (75%) | 10,948 (26%) |
| Diuretics |  |  |  |  |  |  |
| Thiazides/Thiazide-Like | 58,240 (26%) | 21,240 (57%) | 7,616 (23%) | 38,451 (22%) | 12,285 (62%) | 11,859 (28%) |
| Aldosterone Antagonists | 7,179 (3%) | 4,614 (12%) | 706 (2%) | 3,564 (2%) | 2,302 (12%) | 758 (2%) |
| Other (e.g. Potassium sparing, Loop) | 26,759 (12%) | 14,540 (39%) | 2,986 (9%) | 14,230 (8%) | 6,742 (34%) | 4,350 (10%) |
| Others | 17,152 (8%) | 11,125 (30%) | 1,003 (3%) | 7,454 (4%) | 4,831 (24%) | 848 (2%) |
| **Preparation** |  |  |  |  |  |  |
| Any Combination Drug Products | 26,448 (12%) | 9,359 (25%) | 3,509 (11%) | 24,771 (14%) | 7,774 (39%) | 7,673 (18%) |
| **Number of Medications** |  |  |  |  |  |  |
| Average Number of Antihypertensive Medications, mean (St.D.) | 2.1 ± 1.2 | 3.9 ± 0.7 | 1.4 ± 0.5 | 1.9 ± 1 | 3.7 ± 0.6 | 1.4 ± 0.5 |

HTN: hypertension, aTRH: apparent treatment-resistant hypertension.

At the index date, patients with aTRH had a mean BP of 143/79 mm Hg, while those with controlled HTN had a mean BP of 122/74 mm Hg. Among patients with aTRH, only 13,184 (35%) individuals had their BP controlled at the Index date (defined <140/90), compared to 31,154 (95%) in the controlled HTN cohort (Table 3).

**Table 3.** Blood Pressure Control at Index Date for Adults with HTN, aTRH, and stable controlled HTN in the OneFlorida, and REACHnet populations.

| **Population** | **OneFlorida** |  |  | **REACHnet** |  |  |
| --- | --- | --- | --- | --- | --- | --- |
| **BP Characteristic** | **Overall**<br>**N=223,384** | **aTRH**<br>**N=37,274** | **Stable**<br>**Controlled HTN**<br>**N=32,806** | **Overall HTN**<br>**175,229** | **aTRH**<br>**19,863** | **Stable Controlled**<br>**HTN**<br>**42,128** |
| **Average Blood Pressure** |  |  |  |  |  |  |
| Systolic BP, mm Hg, mean (St.D.) | 136 ± 20 | 143 ± 22 | 122 ± 12 | 134 ± 19 | 142 ± 20 | 122 ± 12 |
| Diastolic BP, mm Hg, mean (St. D.) | 79 ± 12 | 79 ± 13 | 74 ± 9 | 78 ± 12 | 78 ± 13 | 74 ± 10 |
| **BP Control** |  |  |  |  |  |  |
| BP Control at Index (<140/90), n (%) | 122,856 (55%) | 13,184 (35%) | 31,154 (95%) | 106,306 (61%) | 7,173 (36%) | 39,574 (94%) |

HTN: hypertension, aTRH: apparent treatment-resistant hypertension, BP: blood pressure.

### aTRH predictors in OneFlorida

Of the 38 independent variables with *P* < 0.05 from the univariate analysis that entered the multivariable stepwise logistic regression model, 24 stayed in the model (Supplemental Tables 3-5). The most significant predictors of aTRH with an odd ratio of ≥1.3 were as follows: black race [odds ratio (OR) 2.81, 95% confidence interval (CI) (2.70-2.92)], CKD stage 4-5 [1.72, (1.58 - 1.89)], BMI ≥25.0 kg/m^2^ [1.65, (1.59 - 1.72)], cardiomegaly [1.53, (1.44 - 1.62)], CKD stage 1-3 [1.48, (1.41 - 1.56)], HFrEF [1.46, (1.34 - 1.58)], HFpEF [1.43, (1.27 - 1.62)], adrenal disorders [1.43, (1.27 - 1.62)], and diabetes [1.42, (1.37 - 1.47)] (Figure 1).

**Figure 1.**
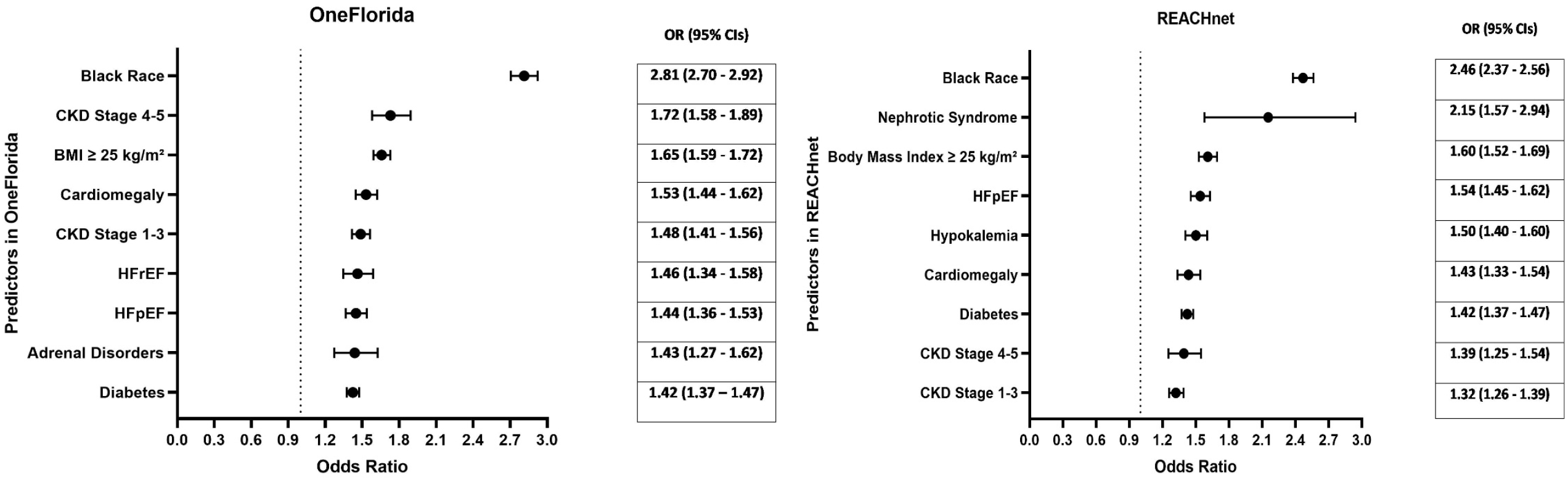
Forest plot of the top aTRH predictors in OneFlorida and REACHnet. HFrEF: heart failure with reduced ejection fraction, HFpEF: heart failure with preserved ejection fraction. CKD: chronic kidney disease, BMI: body mass index. OR: Odds Ratio. CIs: Confidence Intervals

### Prevalence of aTRH and stable controlled HTN, and Patient Characteristics in REACHnet

A total of 241,498 patients within REACHnet were identified with HTN during the study period. Of those, 58,229 patients were excluded for missing BP or drug data. Another 8,040 patients with a diagnosis of secondary HTN or evidence of pregnancy were also excluded, resulting in 175,229 individuals included in the analysis. Among those, 19,863 (11.3%) hypertensive patients met the criteria for aTRH, while 42,128 (24%) patients were identified with stable controlled HTN (Supplemental Figure 2).

Baseline characteristics presented by HTN group are shown in Table 1. Patients with aTRH were younger, and a higher proportion were black and had a BMI ≥25.0 kg/m^2^, compared with the stable controlled HTN group. In addition, at the index date, patients with aTRH had a significantly greater prevalence of diabetes, dyslipidemia, coronary artery disease, stroke, HFpEF, HFrEF, myocardial infarction, CKD (stage 1-3, and stage 4-5), sleep apnea, and other comorbidities, compared to those with stable controlled HTN (Table 1).

### Prescribed Antihypertensive Medications and BP Metrics in REACHnet

The average number of prescribed antihypertensive medications in aTRH and stable controlled HTN patients were 3.7 ± 0.6 and 1.4 ± 0.5, respectively. In patients with aTRH, the most commonly prescribed antihypertensive classes at the index date were BBs (75%), CCBs (74%), and thiazide-like diuretics (62%). In contrast, in those with stable controlled HTN, ACEIs (35%), thiazide-like diuretics (28%), and BBs (26%) were the most commonly prescribed classes (Table 2).

At the index date, patients with aTRH had a mean BP of 142/78 mm Hg, while those with stable controlled HTN had a mean BP of 122/74 mm Hg. 7,173 (36%) individuals in aTRH group had their BP controlled at the index date (defined <140/90), compared to 39,574 (94%) in the stable controlled HTN cohort (Table 3).

### aTRH predictors in REACHnet

Of the 34 independent variables with *P* < 0.05 from the univariate analysis that entered the model, 18 stayed in the model (Supplemental Tables 6-8). The most significant predictors of aTRH with an odd ratio of ≥1.3 in the multivariable stepwise logistic regression model were as follows: black race [odds ratio (OR) 2.46, 95% confidence interval (CI) (2.37-2.56)], nephrotic syndrome [2.15, (1.57 - 2.94)], BMI ≥25.0 kg/m^2^ [1.60, (1.62 - 1.69)], HFrEF [1.54, (1.45 - 1.62)], hypokalemia [1.50, (1.40 - 1.60)], cardiomegaly [1.43, (1.33 - 1.54)], diabetes [1.42, (1.37 - 1.47)], CKD stage 4-5 [1.39, (1.25 - 1.54)], and CKD stage 1-3 [1.32, (1.26 - 1.39)] (Figure 1).

## Discussion

aTRH is a challenging phenotype associated with a high risk of vascular diseases, end organ damage, and poor prognosis [6-8, 10-12]. Our current knowledge of aTRH largely comes from RCTs, closed health systems, and smaller datasets, with less diverse populations [6-8, 10-12, 23]. Accordingly, in this study, we sought to utilize our validated aTRH computable phenotype algorithm to identify the prevalence, characteristics, and predictors of aTRH in large, diverse real-world populations within the OneFlorida, and REACHnet datasets. Our analysis showed that the prevalence of aTRH in OneFlorida was 16.7%, and the prevalence of aTRH in REACHnet was 11.3%. In both populations, aTRH shared similar significant predictors, including black race, diabetes, HFpEF, CKD (stage 4-5 and stage 1-3), cardiomegaly, and BMI ≥25.0 kg/m^2^. Additionally, in both populations aTRH was significantly associated with similar comorbidities when compared to patients with stable controlled HTN. We also observed that the antihypertensive classes prescribed at index date and average BP metrics were comparable in both aTRH populations.

A recent meta-analysis of more than 3 million hypertensive patients reported that the prevalence of aTRH in population-based studies ranged between 4.3% and 29.7% [5]. Our study showed that the prevalence of aTRH in the OneFlorida (16.7%) and REACHnet (11.3%) populations fell within this range. In a prior study, using data from one partner site within OneFlorida (UF Health, 2015-2017) we found a prevalence of aTRH of 15.9%, suggesting consistency in our results and computable phenotype performance [18]. The prevalence estimate of aTRH in the OneFlorida population is comparable to estimates from other studies. For instance, a study performed on Kaiser Permanente Southern California (KPSC) hypertension registry showed 16.9% prevalence of aTRH [24]. Another study reported aTRH prevalence of 17.7% based on an analysis of 2009-2014 data from 4,158 hypertensive patients in the National Health and Nutrition Examination Survey (NHANES) [25]. In addition, results from the Swedish Primary Care Cardiovascular Database (SPCCD) indicated that the rate of aTRH was 17.0% [14]. Likewise, the prevalence of aTRH in the REACHnet population is similar to rates previously reported in the literature. Egan et al. reported that aTRH prevalence was 11.8% among hypertensive patients in NHANES (2005–2008) [26], and the Treating to New Target (TNT) trial also showed that aTRH prevalence was 11.1% [23]. While, the OneFlorida population and the REACHnet population from Ochsner Health have different geographic distributions; OneFlorida including patients in both rural and urban areas, whereas the Ochsner Health population is from more urban areas, the aTRH prevalence is similar in the populations and aligns with prior estimates [5, 25, 26].

The association of aTRH with race and comorbidities is well-recognized. In our study, OneFlorida and REACHnet had a significantly higher proportion of blacks among aTRH patients, compared to stable controlled HTN patients. This finding aligns with the results from the Antihypertensive and Lipid-Lowering Treatment to Prevent Heart Attack Trial (ALLHAT), where blacks were 31% less likely to have their BP controlled [9]. Furthermore, consistent with prior research [6, 8, 14, 15, 27], we observed that aTRH in both populations was significantly associated with a higher prevalence of comorbid conditions like diabetes, dyslipidemia, coronary artery disease, stroke, HFpEF, HFrEF, myocardial infarction, CKD (stage 1-3, and stage 4-5), sleep apnea, and others. While beyond the scope of this study, this association might be explained by the impaired endothelial function and vascular resistance caused by the long-lasting uncontrolled BP [28, 29]. Our results support prior findings that these patients, especially those with CKD and ischemic heart disease, are at high risk of poor prognosis and worse outcomes [6, 16].

Antihypertensive medications are the cornerstone of HTN management. Prior studies found that thiazide diuretics were the most commonly prescribed antihypertensive class as their aTRH definition was based on the use of diuretics along with other antihypertensive classes [15, 30, 31]. On the other hand, we defined aTRH based on the number of antihypertensive classes and BP levels, regardless of diuretics use, therefore, our analysis rather demonstrated that BBs were the most frequently prescribed class in the OneFlorida and REACHnet populations. Our results are in line with those from the TNT trial, NHANES, KPSC, and SPCCD studies [8, 14, 23, 25]. The joint national committee-7 (JNC-7) on prevention, detection, evaluation, and treatment of high BP recommended adding BBs to other antihypertensive classes in case of compelling indications, including HF, post-myocardial infarction, high risk of CVD, and diabetes [32]. Given the high prevalence of these comorbidities in the two populations and the timeframe of our study (2015-2018), during which JNC-7 guidelines were still applied, it seems plausible that BBs were the most commonly prescribed antihypertensive class. However, this is likely to change after the 2017 ACC/AHA clinical practice guidelines recommended adding BBs as a fifth agent after diuretics, CCBs, ACEIs, and ARBs under special circumstances in treating aTRH [33].

Finally, in the multivariable analysis, we observed largely similar significant predictors of aTRH in both populations (black race, raised BMI ≥25.0 kg/m^2,^ diabetes, HFrEF, CKD, and cardiomegaly), suggesting consistency in our results and the populations. These results support findings from previously published studies [6-8, 23]. However, while sex has been previously reported as a risk factor for aTRH [6, 7, 9, 23, 26], sex was not a predictor of aTRH in our analysis. Interestingly, black race was the strongest predictor in both populations, with an odds ratio of more than 2, suggesting a strong association. This correlation could be attributed to the genetic predisposition implicated in renin suppression and salt and water retention in blacks [34]. In addition, our results extend previous findings from the ALLHAT trial that predicted a lower BP control in the presence of diabetes and obesity, possibly explained by insulin resistance and activation of the sympathetic nervous system, respectively [9, 15]. The risk of developing aTRH increases in the presence of CKD [35]. Renal dysfunction increases water and sodium retention, leading to fluid overload and eventually high BP. The Chronic Renal Insufficiency Cohort (CRIC) study reported that aTRH was prevalent in 40.4% of patients with CKD [16]. Unlike other studies, our analysis showed that nephrotic syndrome and hypokalemia are among the significant predictors of aTRH. Ray et al. suggested that nephrotic syndrome increases renal sodium retention, which may explain being a risk factor for aTRH [36]. This increased risk of aTRH in the presence of hypokalemia could be mediated by primary aldosteronism [37]. Overall, our findings suggest that despite the differences between the two populations, aTRH predictors were comparable.

Our study is not without limitations. First, our study did not measure adherence and out-of-office BP to exclude pseudo-resistant or white coat HTN. Therefore, our definition conforms to aTRH. Second, defining aTRH cases was based on the previous BP goal of 140/90 mm Hg. However, since our data were collected before the implementation of the new 2017 ACC/AHA guidelines, when the target BP of 130/80 mm Hg was introduced, we elected to use the 140/90 mm Hg cut-off. Third, patients with HTN and HFrEF included in our study might have received diuretics, renin-angiotensin system (RAS) antagonists, and BBs due to their heart failure, putting them on three antihypertensive medication classes, and increasing their ability to be misclassified as aTRH patients. Finally, although our patient populations are diverse racially and ethnically, representing the U.S general population, our findings may not be generalizable to all populations. Despite these limitations, we used clinical data from clinical practice settings rather than patient-reported data. In addition, our study included large diverse patient populations, reflecting a real-world population. The findings of the discovery and validation populations were largely consistent despite geographical distribution and clinical practice setting differences, demonstrating the interoperability of our computable phenotype.

In conclusion, within the real-world populations of OneFlorida and REACHnet, we demonstrated that the prevalence of aTRH was 16.7% and 11.3%, respectively. Both populations had similar aTRH predictors, antihypertensive prescribing medications at index date, and average BP metrics. Compared to stable controlled HTN, aTRH was significantly associated with similar comorbidities among the two populations. We believe that our results can be utilized to improve healthcare professionals’ understanding of aTRH predictors and associated comorbidities in clinical practice.

## Data Availability

The data included in this study contains PHI, and cannot be readily shared as is. Qualified investigators may apply for data access directly through OneFlorida and REACHnet.

## Non-standard Abbreviations and Acronyms

HTN: Hypertension
ACC/AHA: American College of Cardiology/American Heart Association
RHTN: Resistant Hypertension
aTRH: apparent Treatment-Resistant Hypertension
REACHnet: the Research Action for Health Network
LPHI: Louisiana Public Health Institute
PCORnet: the National Patient-Centered Clinical Research Network
RxCUI: RxNorm Concept Unique Identifiers
BBs: β-Blockers
HFpEF: Heart Failure with preserved Ejection Fraction
HFrEF: Heart Failure with reduced Ejection Fraction
KPSC: Kaiser Permanente Southern California
NHANES: National Health and Nutrition Examination Survey
SPCCD: Swedish Primary Care Cardiovascular Database
TNT: The Treating to New Target
SBP: Systolic Blood Pressure
DBP: Diastolic Blood Pressure
ALLHAT: The Antihypertensive and Lipid-Lowering Treatment to Prevent Heart Attack Trial
JNC=7: The joint national committee-7
CRIC: The Chronic Renal Insufficiency Cohort (CRIC)
RAS: Renin Angiotensin System

## Acknowledgements

The research reported in this publication was conducted in partnership with Research Action for Health Network (REACHnet), funded by the Patient Centered Outcomes Research Institute® (PCORI Award RI-LPHI-01-PS1). REACHnet is a partner network in PCORnet®, which was developed with funding from PCORI®. The content of this publication is solely the responsibility of the authors and does not necessarily represent the views of other organizations participating in, collaborating with, or funding REACHnet or PCORnet®, or of PCORI®.

## Supplemental Material

Figure S1 Flowchart of data preparation in OneFlorida Data Trust population

Figure S2 Flowchart of data preparation in REACHnet population

Tables S1 ICD-9 and 10 codes for comorbidities

Table S2 Antihypertensive classes and medications

Table S3 Univariate analysis in OneFlorida Data Trust

Table S4 Stepwise logistic regression in OneFlorida Data Trust

Table S5 Odds ratio estimates in OneFlorida Data Trust

Table S6 Univariate analysis in REACHnet

Table S7 Stepwise logistic regression in REACHnet

Table S8 Odds ratio estimates in REACHnet

